# Risks and Benefits of Antibiotics vs. COVID-19 Morbidity and Mortality

**DOI:** 10.1101/2020.10.15.20213603

**Authors:** Hisaya Tanioka, Sayaka Tanioka

## Abstract

**Objectives:** The purpose is to analyze the potential association of each antibiotic consumption rate and use ratio with COVID-19 morbidity and mortality, and to investigate the efficacy and safe use of antibiotics against COVID-19.

**Design:** Retrospective statistical analysis study of antibiotic use compared with COVID-19 morbidity and mortality.

**Methods:** Each antibiotic defined daily dose (DDD) per 1000 inhabitants per day as each antibiotic consumption rate was available in the official reports and each antibacterial use ratio data was calculated from them. Coronavirus Disease data were obtained from the WHO Coronavirus Disease Dashboard. The relationships between the sum of DDD, each antibacterial DDD, each antibiotic use ratio, and COVID-19 morbidity and mortality were examined. The statistical correlation was calculated by univariate linear regression analysis and expressed by Pearson’s correlation coefficient.

**Results:** The sum of DDD had no statistical correlation with mortality and morbidity. Cephalosporins were a negative correlation with them. Penicillins had a weak positive correlation with them. Macrolides, quinolones, and sulfonates showed a slightly negative correlation tendency with mortality.

**Conclusions:** Cephalosporins may affect less COVID-19 morbidity and mortality. Penicillins suggest to accelerate them. The combination of cephalosporins with macrolides or quinolones may be a helpful treatment. The difference in antibiotic use between Japan and EU/EEA countries will suggest an explanation for the reduction in morbidity and mortality caused by COVID-19.

## INTRODUCTION

The World Health Organization (WHO) is very clear that antibiotics do not work against viruses but only bacteria, and yet health care providers are using antibiotics in some patients with COVID-19. Patients with viral pneumonia can develop a secondary bacterial infection that may need to be treated with antibiotics, although, this complication is reported to be uncommon early on in the course of COVID-19 pneumonia (1).

On the other hand, some antibiotics have demonstrated several antiviral activities. Azithromycin is a macrolide antibiotic that is being investigated as a potential treatment for people with COVID-19 (2,3,4,5). Clarithromycin suppresses virus growth (5,6). In in vitro laboratory studies, quinolones have demonstrated antiviral activity against human immunodeficiency virus (HIV). And their antiviral activity is identified by inhibition of viral RNA production in a dose-dependent manner (7,8). They also can be considered excellent candidates for the development of anti-Zika virus and anti-chikungunya virus (CHIKV) agents (9). It seems that trimethoprim-sulfamethoxazole suppress human immunodeficiency viral load and provide an excellent clinical response to antiretroviral therapy for HIV (10). Sulfonates can act on Epstein–Barr virus (EBV) and Kaposi sarcoma herpesvirus (11). Antibiotics have been already used for the treatment of community-acquired pneumonia caused by designated, susceptible bacteria, and for the treatment of other bacterial infections in daily practice. Cephalosporins that is more than 60% of them are third-generation, macrolides, and quinolones are frequently used in Japan, and penicillins are frequently used in EU/EEA countries (12,13). The antibiotic use trend is across in East Asia. The morbidity and mortality caused by COVID-19 in Japan and East Asia are less than in EU/EEA countries. Health care providers daily use several antibiotics to treat microbiological infectious patients under COVID-19 status. There are also differences between EU/EEA countries and Japan in how antibiotics are used. Therefore, it may be imagined that each antibiotic daily consumption rate and use ratio may influence COVID-19 morbidity and mortality. If they are associated with morbidity and mortality caused by COVID-19, antibiotics will be suggested to influence COVID-19. Little is known about the potential protective and promotive factors from antibiotics against COVID-19. It is necessary to investigate the protective factors of anti-infective agents that may protect against infection and the factors that improve or promote the outcome if physicians use antibiotics under the condition of COVID-19 epidemic. Therefore, the relationship between each daily antibiotic use and COVID-19 morbidity and mortality should be studied.

This retrospective study aims to determine whether each antibiotic consumption rate and use ratio may have the potential affecting morbidity and mortality caused by COVID-19 and to investigate the effective and safe use of antibiotics against COVID-19.

## METHODS

### Study design

The retrospective statistical analysis study of antibiotic use compared with COVID-19 morbidity and mortality.

### Setting and samples

This single-center study was obtained by the institutional ethics review board, and this study was the retrospective cohort study using public data. Data were collected for antibiotic consumption rate and use ratio, and COVID-19 morbidity and mortality.

### Procedure

The antibiotic consumption data from the annual epidemiological report for 2019 that contained the consumption of antibacterials for systemic use by EU/EEA countries in 2018 expressed as DDD per 1000 inhabitants per day was used (12). Japan data was obtained from the antimicrobial selling data in 2019 (13). That is, these data represented each antibiotic consumption rate. Each antibacterial use ratio was calculated from these data. COVID-19 morbidity per 0.1 M population and mortality caused by this disease per 0.1 M population in each of the countries were obtained from the WHO coronavirus disease (COVID-19) Dashboard on 25 May 2020. Table 1 showed the population, morbidity per 0.1 M population, mortality per 0.1 M population, the sum of consumption as DDD per 1000 inhabitants per day, and each antibiotic consumption as DDD per 1000 inhabitants per day (tetracyclines: J01A, macrolides: J01F, cephalosporins: J01D, penicillins: J01C, quinolones: J01M, and sulfonamides: J01E) by each of the countries. Table 2 showed morbidity per 0.1 M population with each number, mortality per 0.1 M population with each number, and each antibiotic use ratio by each of the countries.

**Table 1.** DDD, Each Antibacterial DDD, COVID-19 Morbidity and Mortality in EU/EEA and Japan.

| Nation | Population<br>per 0.1 M | Morbidity<br>per 0.1M | Mortality<br>per 0.1M | DDD<br>/1000 | Each Antibacterial DDD per 1000 |  |  |  |  |  |
| --- | --- | --- | --- | --- | --- | --- | --- | --- | --- | --- |
|  |  |  |  |  | Tet | Mac | Ceph | Pen | Qui | Sul |
| Greece | 107.3 | 26.47 | 1.55 | 32.4 | 2.9 | 6.3 | 7.9 | 11.1 | 2.7 | 0.3 |
| Romania | 194.7 | 88.29 | 5.78 | 25.0 | 0.9 | 3.0 | 5.1 | 11.3 | 2.1 | 0.8 |
| Belgium | 114.3 | 488.11 | 79.68 | 20.8 | 1.9 | 4.0 | 1.3 | 10.1 | 1.2 | 0.2 |
| Cyprus | 11.9 | 77.14 | 2.02 | 31.1 | 3.6 | 3.3 | 5.3 | 13.3 | 4.6 | 0.2 |
| France | 669.8 | 214.13 | 41.81 | 23.6 | 3.1 | 2.9 | 1.4 | 13.3 | 1.3 | 0.4 |
| Luxembourg | 6.1 | 648.85 | 17.95 | 20.7 | 2.1 | 3.5 | 2.9 | 8.5 | 2.0 | 0.3 |
| Italy | 604.8 | 374.83 | 53.09 | 19.5 | 0.5 | 3.9 | 2.1 | 8.9 | 2.7 | 0.7 |
| Ireland | 48.7 | 497.97 | 32.05 | 20.9 | 2.6 | 4.1 | 0.6 | 9.8 | 0.8 | 1.0 |
| Portugal | 102.8 | 286.30 | 12.13 | 17.2 | 0.9 | 2.7 | 1.6 | 9.2 | 1.2 | 0.3 |
| Malta | 4.8 | 117.56 | 1.24 | 22.8 | 1.5 | 4.5 | 2.7 | 6.3 | 2.3 | 0.3 |
| Iceland | 3.5 | 511.93 | 2.84 | 20.4 | 5.0 | 1.6 | 0.6 | 8.9 | 0.8 | 2.5 |
| Croatia | 40.9 | 54.57 | 2.35 | 17.0 | 1.0 | 2.8 | 2.5 | 7.8 | 1.5 | 0.5 |
| Spain | 468.0 | 420.85 | 61.44 | 24.3 | 1.5 | 3.0 | 2.3 | 13.7 | 2.7 | 0.4 |
| UK | 656.4 | 395.35 | 56.00 | 16.3 | 4.6 | 2.6 | 0.2 | 6.2 | 0.6 | 0.8 |
| Slovakia | 54.5 | 27.43 | 0.51 | 20.5 | 1.7 | 4.7 | 6.2 | 5.0 | 2.1 | 0.4 |
| Poland | 379.7 | 50.75 | 2.50 | 23.0 | 2.3 | 6.0 | 3.0 | 6.8 | 1.5 | 0.5 |
| Finland | 55.2 | 115.92 | 5.45 | 13.2 | 3.1 | 0.7 | 1.9 | 4.4 | 0.6 | 1.0 |
| Bulgaria | 70.3 | 32.13 | 1.59 | 19.4 | 1.6 | 4.0 | 4.4 | 5.5 | 2.8 | 0.9 |
| Czech ep. | 106.3 | 81.35 | 2.84 | 19.5 | 2.0 | 4.0 | 2.2 | 8.3 | 0.9 | 0.9 |
| Norway | 52.3 | 158.07 | 4.46 | 14.0 | 2.6 | 0.9 | 0.1 | 5.5 | 0.3 | 0.7 |
| Denmark | 57.0 | 193.75 | 9.67 | 13.7 | 1.4 | 1.5 | 0.0 | 9.2 | 0.4 | 0.7 |
| Lithuania | 28.0 | 55.79 | 2.14 | 15.4 | 1.4 | 2.2 | 2.9 | 8.5 | 2.0 | 0.3 |
| Germany | 829.1 | 214.48 | 9.88 | 11.9 | 1.6 | 1.9 | 2.5 | 3.7 | 1.0 | 0.6 |
| Slovenia | 20.7 | 70.87 | 5.02 | 11.7 | 0.5 | 1.8 | 0.3 | 6.9 | 0.4 | 0.7 |
| Sweden | 101.8 | 298.40 | 36.33 | 10.8 | 2.2 | 0.3 | 0.1 | 5.7 | 0.5 | 0.3 |
| Austria | 87.9 | 185.12 | 7.19 | 10.4 | 0.4 | 2.3 | 1.4 | 4.8 | 1.0 | 0.2 |
| Hungary | 97.8 | 36.79 | 4.81 | 13.7 | 1.1 | 2.3 | 2.1 | 4.6 | 2.3 | 0.1 |
| Latvia | 19.3 | 52.44 | 1.09 | 11.4 | 2.0 | 2.2 | 0.6 | 4.4 | 1.1 | 0.7 |
| Estonia | 13.2 | 135.68 | 4.85 | 10.2 | 1.2 | 2.3 | 1.2 | 3.7 | 0.8 | 0.4 |
| Netherlands | 172.3 | 256.81 | 33.16 | 8.9 | 1.9 | 1.4 | 0.0 | 2.9 | 0.7 | 0.4 |
| Japan | 1261.8 | 12.99 | 0.61 | 13.3 | 0.8 | 3.9 | 3.8 | 1.5 | 2.4 | 0.7 |

**Table 2.** Each Antibacterial Use Ratio, COVID-19 Morbidity and Mortality in EU/EEA and Japan.

| Nation | Morbidity |  | Mortality |  | Each Antibacterial Use Ratio (%) |  |  |  |  |  |
| --- | --- | --- | --- | --- | --- | --- | --- | --- | --- | --- |
|  | / 0.1M (number) |  | / 0.1M (number) |  | Tet | Mac | Ceph | Pen | Qui | Sul |
| Greece | 26.47 | (2840) | 1.55 | (166) | 9.0 | 19.4 | 24.4 | 34.3 | 8.3 | 0.9 |
| Romania | 88.29 | (17191) | 5.78 | (1126) | 3.6 | 12.0 | 20.4 | 45.2 | 8.4 | 3.2 |
| Belgium | 488.11 | (55791) | 79.68 | (9108) | 9.1 | 14.4 | 6.3 | 48.6 | 5.8 | 1.0 |
| Cyprus | 77.14 | (918) | 2.02 | (24) | 11.6 | 10.6 | 17.0 | 42.8 | 14.8 | 0.6 |
| France | 214.13 | (143427) | 41.81 | (28002) | 13.1 | 12.3 | 5.9 | 56.4 | 5.5 | 1.7 |
| Luxembourg | 648.85 | (3958) | 17.95 | (109) | 10.1 | 16.9 | 14.0 | 41.0 | 9.7 | 1.4 |
| Italy | 374.83 | (226699) | 53.09 | (32169) | 2.6 | 20.0 | 10.8 | 45.6 | 13.8 | 3.6 |
| Ireland | 497.97 | (24251) | 32.05 | (1561) | 12.4 | 19.6 | 2.9 | 46.9 | 3.8 | 4.8 |
| Portugal | 286.30 | (29432) | 12.13 | (1247) | 5.2 | 15.7 | 9.3 | 53.5 | 7.0 | 1.7 |
| Malta | 117.56 | (569) | 1.24 | (6) | 6.6 | 19.7 | 11.8 | 27.6 | 10.1 | 1.3 |
| Iceland | 511.93 | (1802) | 2.84 | (10) | 24.5 | 7.8 | 2.9 | 43.6 | 3.9 | 12.3 |
| Croatia | 54.57 | (2232) | 2.35 | (96) | 5.9 | 16.5 | 14.7 | 45.9 | 8.8 | 2.9 |
| Spain | 420.85 | (196958) | 61.44 | (28752) | 6.2 | 12.3 | 9.5 | 56.4 | 11.1 | 1.6 |
| UK | 395.35 | (259509) | 56.00 | (36758) | 28.2 | 16.0 | 1.2 | 38.0 | 3.7 | 4.9 |
| Slovakia | 27.43 | (1495) | 0.51 | (28) | 8.3 | 22.9 | 30.2 | 24.4 | 10.2 | 2.0 |
| Poland | 50.75 | (19268) | 2.50 | (948) | 10.0 | 26.1 | 13.0 | 29.6 | 6.5 | 2.2 |
| Finland | 115.92 | (6399) | 5.45 | (301) | 23.5 | 5.3 | 14.4 | 33.3 | 4.5 | 7.6 |
| Bulgaria | 32.13 | (2259) | 1.59 | (112) | 8.2 | 20.6 | 22.7 | 28.4 | 14.4 | 4.6 |
| Czech ep. | 81.35 | (8647) | 2.84 | (302) | 10.3 | 20.5 | 11.3 | 42.6 | 4.6 | 4.6 |
| Norway | 158.07 | (8267) | 4.46 | (233) | 18.6 | 6.4 | 0.7 | 39.3 | 2.1 | 5.0 |
| Denmark | 193.75 | (11044) | 9.67 | (551) | 10.2 | 10.9 | 0 | 67.2 | 2.9 | 5.1 |
| Lithuania | 55.79 | (1562) | 2.14 | (60) | 9.1 | 14.3 | 18.8 | 55.2 | 13.0 | 1.9 |
| Germany | 214.48 | (177827) | 9.88 | (8198) | 13.4 | 16.0 | 21.0 | 31.1 | 8.4 | 5.0 |
| Slovenia | 70.87 | (1467) | 5.02 | (104) | 4.3 | 15.4 | 2.6 | 59.0 | 3.4 | 6.0 |
| Sweden | 298.40 | (30377) | 36.33 | (3698) | 20.4 | 2.8 | 0.9 | 52.8 | 4.6 | 2.8 |
| Austria | 185.12 | (16272) | 7.19 | (632) | 3.8 | 22.1 | 13.5 | 46.2 | 9.6 | 1.9 |
| Hungary | 36.79 | (3598) | 4.81 | (470) | 8.0 | 16.8 | 15.3 | 33.6 | 16.8 | 0.7 |
| Latvia | 52.44 | (1012) | 1.09 | (21) | 17.5 | 19.3 | 5.3 | 38.6 | 9.6 | 6.1 |
| Estonia | 135.68 | (1791) | 4.85 | (64) | 11.8 | 22.5 | 11.8 | 36.3 | 7.8 | 1.9 |
| Netherlands | 256.81 | (44249) | 33.16 | (5714) | 21.3 | 15.7 | 0 | 32.6 | 7.9 | 4.5 |
| Japan | 12.99 | (16395) | 0.61 | (773) | 6.0 | 29.3 | 28.6 | 11.3 | 18.0 | 5.3 |

### Statistical analysis

Statistical analyses were carried out using univariate linear regression analysis and Pearson’s correlation coefficient. A p-value of < 0.05 was regarded as significant. Data were analyzed statistical software available through Microsoft Excel 2013 (Microsoft Corporation Redmond, Washington). We analyzed two kinds of relationships. One was the relationship between each antibiotic DDD per 1000 inhabitants per day and COVID-19 morbidity and mortality. The other was the relationship between each antibiotic use ratio and them. These analyses were examined by tetracyclines (J01A), macrolides (J01F), cephalosporins (J01D), penicillins (J01C), quinolones (J01M), and sulfonates (J01E).

## RESULTS

Morbidity per 0.1 M population was 199.39±175.39 (n=31) and mortality per 0.1 M population was 16.83±22.65 (n=31).

The results of the two analyses were calculated by univariate linear regression analysis and expressed by Pearson’s correlation coefficient. The regression line represented as Y: morbidity or mortality, and X: antibacterial consumption rate or use ratio.

### The statistical analysis between antibacterial consumption rate and COVID-19 morbidity and mortality

1. **Defined Daily Dose (DDD):** The sum was a mean±SD: 17.84±5.98. The sum of DDD had no relationship with morbidity (Y = 1.64X+161.92; r = 0.05; significance: NS; p = 0.77) and mortality (Y = 0.27X+11.34; r = 0.07; significance: NS; p = 0.69).
2. **Macrolides (J01F):** The consumption rate was a mean±SD: 2.92±1.42. There was no relationship with morbidity (Y = −16.03X+246.25; r = −0.13; significance: NS; p = 0.49) and with mortality (Y = −0.36X+11.41; r = −0.02; significance: NS; p = 0.90).
3. **Cephalosporins (J01D):** The consumption rate was a mean±SD: 2.23±1.94. A negative correlation with morbidity (Y = −38.43X+285.18; r=-0.43; significance: p < 0.05; p = 0.02) and mortality (Y = −3.89X+24.89; r = −0.50; significance: p<0.01; p = 0.004) was seen.
4. **Quinolones (J01M):** The consumption rate was (a mean±SD: 1.53±1.42). There was a negative weak correlation with morbidity (Y = −39.20X+259.20; r = −0.22; significance: NS; p = 0.24) and no relationship with mortality (Y = −2.09X+19.39; r = −0.10; significance: NS; p = 0.61).
5. **Sulfonates and trimethoprim (J01E):** The consumption rate was a mean±SD: 0.59±0.44. There was a weak positive correlation with morbidity (Y = 95.03X+145.60; r = 0.24; significance: NS; p = 0.20) and no relationship with mortality (Y = −7.02X+20.32; r = −0.14; significance: NS; p = 0.45) was seen.
6. **Tetracyclines (J01A):** The consumption rate was a mean±SD: 1.93±1.11. There was a week positive correlation with morbidity (Y = 49.98X+10.97; r = 0.32; significance: NS; p = 0.08) and no relationship with mortality (Y = 2.48X+11.41; r = −0.13; significance: NS; p = 0.49).
7. **Penicillins (J01C):** The consumption rate was a mean±SD: 7.41±3.16. A week positive correlation with morbidity (Y = 16.97X+73.57; r = 0.31; significance: NS; p = 0.09) and with mortality (Y = 2.34X-1.17; r = 0.34; significance: NS; p = 0.06) was seen.

### The statistical analysis between antibacterial use ratio and COVID-19 morbidity and mortality

1. **Macrolides (J01F):** The use ratio was a mean±SD: 16.40±1.42. A negative weak correlation with morbidity (Y = −9.20X+350.37; r = −0.31; significance: NS; p = 0.09) and with mortality (Y = −0.74X+28.13; r = −0.20; significance: NS; p = 0.27) was seen.
2. **Cephalosporins (J01D):** The use ratio was a mean±SD: 11.65±8.49. A negative correlation with morbidity (Y = −0.30X+319.43; r = −0.50; significance: p< 0.01; p = 0.00) and mortality (Y = −1.12X+29.27; r = −0.44; significance: p<0.05; p = 0.01) was found.
3. **Quinolones (J01M):** The use ratio was a mean±SD: 8.36±4.21. There was a negative weak correlation with morbidity (Y = −13.44X+311.64; r = −0.32; significance NS; p = 0.08) and no relationship with mortality (Y = −0.90X+23.71; r = −0.18; significance: NS; p = 0.35).
4. **Sulfonates and trimethoprim (J01E):** The use ratio was a mean±SD: 3.58±2.47. There was no relationship with morbidity (Y = 8.97X+167.23; r = 0.13; significance: NS; p = 0.50) and with mortality (Y= −1.63X+22.07; r = −0.19; significance: NS; p = 0.32) was seen.
5. **Tetracyclines (J01A):** The use ratio was a mean±SD: 11.38±6.64. There was a week positive correlation with morbidity (Y = 6.99X+119.86; r = 0.28; significance: NS; p = 0.08) and no relationship with mortality (Y = 0.45X+11.07; r = 0.14; significance: NS; p = 0.47).
6. **Penicillins (J01C):** The use ratio was a mean±SD: 41.53±11.78. A weak positive correlation with morbidity (Y = 11.21X+5.07; r = 0.34; significance: NS; p = 0.06) and with mortality (Y = 0.64X-10.26; r = 0.35; significance: NS; p = 0.06) was revealed.

## DISCUSSION

### Principle findings

The antibiotic consumption dose for systemic use by EU/EEA countries in 2018 was expressed as DDD per 1000 inhabitants per day. Therefore, the relationship between each DDD per 1000 inhabitants per day, and COVID-19 morbidity and mortality represented whether each antibiotic consumption rate is related to COVID-19. The relationship between the antibiotic use ratio and COVID-19 morbidity and mortality indicated whether the antibiotic use trend affects COVID-19. The sum of DDD suggests no affect morbidity and mortality caused by COVID-19. However, each antibiotic analysis represents various analytical results. Cephalosporins suggest decreasing morbidity and mortality caused by COVID-19. Penicillins may have the potential to accelerate them. Quinolones suggest the potential to reduce morbidity, and the weak potential against mortality. Macrolides may have the potential for reducing morbidity and mortality. Therefore, macrolides and quinolones may suggest working against COVID-19. Sulfonates and trimethoprim have almost the same mortality affect as quinolones. However, they may suggest increasing morbidity. Tetracyclines may increase morbidity, but may not affect mortality. The results of these analyses were similar to those of EU/EEA countries alone. Therefore, cephalosporins, macrolides, and quinolones may be potentially effective against COVID-19 (SARS-CoV-2). And, sulfonates and trimethoprim may have the potential reducing mortality.

Cephalosporins, macrolides, and quinolones are frequently used in Japan, but penicillins are frequently used in European countries (12,13). The different antibiotic use trends may suggest an explanation for less morbidity and mortality caused by COVID-19 in Japan rather than in EU/EEA countries. The antibiotic use trend in Japan is across East Asia. Although the analytical data is from 2018 and is not the latest, the antibiotic consumption rate and use trend from the previous report are not so different. Therefore, the results of this study will appear to fit in with the latest.

### Penicillins

The results imply that it is prudent to avoid penicillins use in the current status of the COVID-19 epidemic, as penicillins may suggest increasing morbidity and mortality caused by COVID-19. Penicillins may appear to create an environment for COVID-19 proliferation or may help the virus spread rapidly through cells.

### Macrolides

Macrolides have been shown to be active in vitro against RNA viruses. Azithromycin is thought to have antiviral and anti-inflammatory activity and may work synergistically with other antiviral treatments. Azithromycin has demonstrated antiviral activity against Zika virus and against rhinoviruses from in vitro studies. Azithromycin is a macrolide antibiotic that is being investigated as a potential treatment for people with COVID-19 caused by the new coronavirus (SARS-CoV-2) (2,3,4,5). Clarithromycin decreases interferon (IFN)-γ and increases IL-10 levels. Clarithromycin promotes the expansion of immunosuppressive CD11b+Gr-1+ cells essential for the immunomodulatory properties of macrolides (6). Clarithromycin suppresses virus growth (5,6). Ivermectin is a cyclic lactone oral anthelmintic that belongs to the macrolides group. Ivermectin is anti-parasitic previously shown to have broad-spectrum antiviral activity in vitro, and it is an inhibitor of SARS-CoV-2 (14). Indeed, it seems that the mortality rate caused by COVID-19 is low in Onchocerciasis endemic areas of Africa (1). However, from an action point of view, these drugs seem to be effective only in the early stages.

### Quinolones

Quinolone derivatives have been shown to inhibit human immunodeficiency virus (HIV) replication at the transcriptional level (7). Richter et al. reported that introducing an aryl group into the piperazine moiety of fluoroquinolone changed its activity from antibacterial to antiviral, with specific effects on human immunodeficiency virus (HIV) (8). Antiviral activity is confirmed by dose-dependently inhibiting viral RNA production (9).

### Sulfonates and trimethoprim

Sulfonates and trimethoprim show almost the same mortality effect as quinolones. So, they may work against COVID-19 mortality. Trimethoprim-sulfamethoxazole appears to be effective in suppressing human immunodeficiency virus (HIV) load and antiretroviral therapy (10). It has antimalarial and antibacterial properties. Since trimethoprim-sulfamethoxazole is used in the current state of COVID-19, its function against RNA viruses may be due to its antimalarial properties. Because, in principle, antibacterial agents do not act on viruses. Sulfonates can also act on herpesvirus such as EBV and Kaposi sarcoma herpesvirus belonging to DNA virus (11). Hence, antimalarial properties may act on RNA and DNA viruses.

### Cephalosporins

Strangely, in this study, the higher cephalosporins consumption rate and use ratio will make lower morbidity and mortality caused by COVID-19 than other antibacterials. Cephalosporins have not been proven effective against viruses. β-lactam antibiotics including cephalosporins have a bacteriostatic effect. The bacteriostatic action means the action of suppressing the growth of bacteria. β-lactam antibiotics act by inhibiting peptidoglycan synthase, which is an enzyme required for synthesizing bacterial cell walls. However, cephalosporins suggest inhibiting SARS-CoV-2.

There are four effective cases of cephalosporins (Ceftriaxone) combination therapy with minocycline (broad-spectrum tetracycline) in Japan (15). Tetracyclines do not affect mortality caused by COVID-19 from this study. Therefore, the results of this study and these effective cases imply that cephalosporins may work against SARS-CoV-2.

### Based on empiric treatment for patients with MERS coronavirus (MERS-CoV)

According to the previous report (16) for patients with MERS coronavirus (MERS-CoV) that is the same coronavirus of SARS-CoV-2, empirical treatment with neuraminidase inhibitors and an association of antibiotics effective against S. pneumoniae and L. pneumophila are the key management of hospitalized patients. Third-generation cephalosporins are effective against S. pneumoniae, and macrolides and quinolones are effective against L. pneumophila. Macrolides (azithromycin or clarithromycin) or quinolones (levofloxacin or moxifloxacin) are the standard treatment for Legionella pneumonia in humans, with levofloxacin being considered first-line with increasing resistance to azithromycin. The results of this study show the similarities in the use of antibiotic agents in the conclusions of the previous report (16). The antibiotic combination of cephalosporins with trimethoprim-sulfamethoxazole (TS) may also be an effective therapy from the perspective of reducing COVID-19 mortality.

### Implications for practice

When a physician needs to treat several secondary bacterial infections that two different antibiotic combinations are often used, the combination therapy with cephalosporins and quinolones or macrolides may be an effective treatment under the condition of COVID-19 epidemic period. In general, there are no specific antiviral drugs or vaccines for SARS-CoV-2. All of the drug treatment options come from experience treating SARS, MERS, or some other new influenza virus previously. Active symptomatic support continues to be the key to treatment. This retrospective study suggests that the treatment regimen proposed by Bleibtreu et al. (16) may be effective for COVID-19. However, the efficacy of this regimen needs to be further confirmed.

## Conclusions

The results of this study imply that penicillins should be avoided to use under the condition in COVID-19. Empirical treatment with neuraminidase inhibitors and the combination of cephalosporins and macrolides or quinolones are suggested to be an effective treatment for COVID-19.

## Data Availability

All data generated or analyzed during this study are included in this article.

https://www.ecdc.europa.eu/sites/default/files/documents/Antimicrobial-consumption-EU-EEA.pdf

http://amrcrc.ncgm.go.jp/surveillance/020/sales_2020.pdf

## Abbreviations

DDD: Defined Daily Dose
WHO: World Health Organization
COVID-19: Coronavirus Disease 2019
HIV: Human Immunodeficiency Virus
EBV: Epstein–Barr virus
CHIKV: Chikungunya virus
SARS-CoV-2: Severe Acute Respiratory Syndrome Coronavirus-2
MERS: Middle East respiratory syndrome
J01A: Tet: Tetracyclines
J01C: Pen: Penicillins
J01D: Ceph: Cephalosporins and other beta-lactams
J01E: Sul: Sulfonates and trimethoprim
J01F: Mac: Macrolides, lincosamides, and streptogramins
J01M: Qui: Quinolones

## Declaration

### Compliance with ethical standards

We declare that we have no conflict of interest.

### Ethical statement

This retrospective study complies with ethical standards.

### Availability of data and materials

All data generated or analyzed during this study are included in this article.

### Author contribution

This study was done by ourselves. We carried out the design of the study, performed the statistical analysis, and prepared the manuscript. HT wrote the final manuscript.

